# Clinical characteristics and outcomes of adult patients admitted with COVID-19 in East London: a retrospective cohort analysis

**DOI:** 10.1101/2020.10.08.20193623

**Authors:** Daryl Oswald Cheng, Claire Jacqueline Calderwood, Erik Wilhelm Skyllberg, Adam Denis Jeremy Ainley

**Affiliations:** Barking, Havering and Redbridge University Hospitals NHS Trust, London, UK

## Abstract

**Background:** Descriptions of clinical characteristics of patients hospitalised with coronavirus disease 2019 (COVID-19), their clinical course and short-term in- and outpatient outcomes in deprived urban populations in the United Kingdom are still relatively sparse. We describe the epidemiology, clinical course, experience of non-invasive ventilation and intensive care, mortality and short-term sequalae of patients admitted to two large District General Hospitals across a large East London NHS Trust during the first wave of the pandemic.

**Methods:** A retrospective analysis was carried out on a cohort of 1,946 patients with a clinical or laboratory diagnosis of COVID-19, including descriptive statistics and survival analysis. A more detailed analysis was undertaken of a subset of patients admitted across three Respiratory Units in the trust.

**Results:** Increasing age, male sex and Asian ethnicity were associated with worse outcomes. Increasing severity of chest X-ray abnormalities trended with mortality. Radiological changes persisted in over 50% of cases at early follow up (6 weeks). Ongoing symptoms including hair loss, memory impairment, breathlessness, cough and fatigue were reported in 67% of survivors, with 42% of patients unable to return to work due to ongoing symptoms.

**Conclusions:** Understanding the acute clinical features, course of illness and outcomes of COVID-19 will be vital in preparing for further peaks of the pandemic. Our initial follow up data suggest there are ongoing sequalae of COVID-19 including persistent symptoms and radiological abnormalities. Further data, including longer term follow up data, are necessary to improve our understanding of this novel pathogen and associated disease.

**Section 1: What is already known on this topic:** Previous studies have reported that increasing age, male sex, Black and Asian ethnicity increased risk of death for patients admitted to hospital with coronavirus disease 2019 (COVID-19). There is little published literature regarding the follow up of patients with COVID-19.

**Section 2: What this study adds:** Our study is one of the first with follow up data for patients admitted to hospital with COVID-19. We show that radiological abnormality persisted at 6 weeks in over 50% of patients, as well as significantly increased breathlessness in patients without baseline dyspnoea. Our study confirms that increasing age, male sex and Asian ethnicity increased risk of death for patients, but also in an ethnically and socioeconomically diverse population in East London.

## 1. Introduction

### 1.1 Background and Rationale

Coronavirus disease 2019 (COVID-19) is a respiratory illness caused by the novel coronavirus, severe acute respiratory syndrome coronavirus 2 (SARS-CoV-2). In March 2020 it was designated a pandemic by the World Health Organization; at the time of writing there have been more than 20 million cases worldwide and 750,000 deaths associated with COVID-19.^1,2^ Within Europe, the UK has experienced a high burden of COVID-19 with more than 300,000 cases, the second highest number of total cases in the region, and more than 46,000 deaths.^1^

London has the highest age-standardised mortality rate from COVID-19 in the UK (85.7 deaths per 100,000 individuals).^3^ It is the most ethnically diverse urban area of the UK. The population which Barking, Havering and Redbridge University NHS Hospitals Trust (BHRUT) serves is similarly diverse and includes boroughs with higher indices of multiple deprivation.^4^ The population is also one of the largest in London served by a single NHS Trust. At the time of writing (15 August 2020) 3,030 individuals have been diagnosed with suspected or confirmed COVID-19 from Upper Tier Local Authorities (UTLAs) under the BHRUT catchment.^9^

Ethnicity is one of the major public health questions surrounding COVID-19,^5^ with evidence that Black, Asian and minority ethnic (BAME) groups are disproportionately represented in intensive care admissions^6^ and have higher mortality, even after adjusting for geographic, socio-economic,^7^ cardiometabolic and behavioural factors.^8^

### 1.2 Objectives

The reporting of our local experience forms an important comparison to that of other large hospital trusts in populous urban centres nationally and internationally. The objectives were to describe the population of individuals admitted to BHRUT with COVID-19 between 1 March 2020 and 8 June 2020 including in-hospital mortality and early outpatient outcomes; and to explore factors which may be associated with increased mortality within this population.

Additionally, we present an analysis of a nested cohort of patients admitted to the Respiratory Units (RU). The RUs comprise a total of 90 beds across two sites and admitted the first patients with COVID-19, before rapidly evolving to preferentially admit patients with more severe disease including those requiring non-invasive ventilation (NIV), which was delivered on the Unit. We describe the demographic, clinical, laboratory and radiological findings and disease course and clinical outcomes of these patients.

## 2 Methods

### 2.1 Design and participants

This retrospective cohort study included adult patients (age ≥18 years old) admitted across two hospitals that comprise a large NHS Trust in London, UK with a clinical or laboratory diagnosis of COVID-19, from 1 March 2020 to 8 June 2020. The reporting of this study is in accordance with the guidelines set out in the STROBE (STrengthening the Reporting of OBservational studies in Epidemiology) statement. Additional data were collected for a subset of patients admitted to the RUs between 10 March 2020 (the date of admission of the first patient with COVID-19) and 26 April 2020, included here as a nested cohort.

Follow up data for patients discharged following a diagnosis COVID-19 are included as they are available at the time of writing. All patients discharged after a severe illness with COVID-19 are being followed up by these clinics, with the first patients being those identified as admitted to the RU.

### 2.2 Data collection

Data were collected from a combination of electronic health records of vital signs and coded admission databases. Data extracted included age, sex, ethnicity, length of stay, and discharge outcome. For the nested RU dataset, these electronic datasets were combined with data collected from manual review of electronic and paper-based patient records. Data collected included patient demographics, clinical, laboratory and radiological findings, progress during admission including the requirement for organ support, and outcome. All data were linked using unique patient identifiers before subsequent anonymisation for analysis.

Follow up data were collected from clinic records for patients attending for follow up at approximately 6 weeks from their discharge using locally adapted British Thoracic Society guidelines.^10^ Data collected included patient demographics, Medical Research Council (MRC) breathlessness scales at baseline (evaluated retrospectively) and present; results of screening tools for anxiety (Generalised Anxiety Disorder scale-2 [GAD-2], depression (Patient Health Questionnaire-2 [PHQ-2]) and post-traumatic stress disorder (Trauma Screening Questionnaire [TSQ]); functional impairment and return to work status. Chest radiographs (CXR) were reviewed from admission and post-discharge.

### 2.3 Analysis

Analysis was performed using the R statistical computing environment version 4.0.1.^13,14^ For continuous variables, analysis of variance tests was used. For categorical variables, Pearson’s Chi-square tests or Fischer’s exact tests were used. Cox proportional hazard models were used for survival analyses, and the proportional hazard assumptions were checked using Schoenfeld residuals. Complete case records analysis was used, with missing data assumed to be so at random.

### 2.4 Definitions

COVID-19 cases were defined as either a laboratory confirmed or clinically suspected diagnosis of COVID-19. Patients who tested positive via real-time polymerase chain reaction for SARS-CoV-2 in a respiratory tract sample were classed as laboratory confirmed. A clinically suspected case was defined as a patient who tested negative for COVID-19, but fit clinical and radiological definitions for diagnosis of COVID-19 from Public Health England (PHE).^11^ Treatment escalation definitions are: level 1 is for maximal therapy delivered in a medical ward environment; level 2 is for single organ support in a higher dependency area including the RU; and level 3 for multi-organ support including invasive mechanical ventilation in an intensive care setting. Ethnicity definitions were those defined by the UK Office for National Statistics.^4^ Obesity is defined as a recorded body mass index of ≥30 kg/m^2^. Frailty was recorded as the Rockwood Clinical Frailty Score (CFS). An NIV trial is defined as any time on either continuous positive airway pressure (CPAP) or bilevel positive airway pressure (BiPAP). CXR classification and severity was assessed by a member of the respiratory team according to the British Society of Thoracic Imaging (BSTI) classification.^12^

### 2.5 Ethics

These analyses were completed as part of ongoing service evaluation in order to facilitate future planning and ongoing service requirements for patients with COVID-19, in collaboration with the Trust Research and Development Team. The collection of clinical audit data was prospectively registered with the Trust Clinical Audit Department prior to data collection. In line with the NHS Health Research Authority guidance, Research Ethics Committee approval was not required, and the Caldicott Guardian was consulted for approval for the use of anonymised patient data.

### 2.6 Patient and public involvement

Due to the retrospective nature of the study, patients and the public were not involved prior to the collection of data, study design or recruitment to the study. Follow up data fields were adapted from BTS guidelines, which has patient and public involvement in the form of Lay Trustees. We intend to disseminate the main results of the study using the Trusts’ public engagement channels.

## 3. Results

### 3.1 Overall cohort

#### 3.1.1. Demographics and ethnicity

Data were available for 2,091 admissions, representing 1,946 patients, with a coded diagnosis of suspected or confirmed COVID-19, admitted from 1 March 2020 to 8 June 2020. Median age was 73 years (interquartile range [IQR] 57 – 84 years); 42.1% patients were female. Of 1,781 patients with ethnicity data available, 1,250 (70.2%) were of White ethnicity, 23 (1.3%) Mixed, 313 (17.6%) Asian, 154 (8.6%) Black, 41 (2.3%) Other. (Table 1)

**Table 1.** Distribution of socio-demographic factors for patients admitted to the trust with COVID-19, deaths per person-years at risk, and adjusted hazard ratios from Cox regression model

|  | Total | Deaths | aHR | 95% CI | p |
| --- | --- | --- | --- | --- | --- |
| Overall | 1946 | 594 | - | - | - |
| Age / years (n = 1855) |  |  |  |  | <0.001 |
| Median (IQR) | 73 (57-84) | 81 (72-87) | - | - |  |
| <60 | 560 (30.2%) | 51 (9.5%) | Ref | - |  |
| 60-80 | 666 (35.9%) | 206 (38.3%) | 2.95 | 2.12-4.11 |  |
| >80 | 629 (33.9%) | 281 (52.2%) | 4.44 | 3.19-6.19 |  |
| Gender (n = 1855) |  |  |  |  | 0.004 |
| Female | 820 (44.2%) | 205 (38.1%) | Ref | - |  |
| Male | 1035 (55.8%) | 333 (61.9%) | 1.30 | 1.09-1.56 |  |
| Ethnicity (n = 1781) |  |  |  |  | 0.1 |
| White | 1250 (70.2%) | 392 (76.6%) | Ref | - |  |
| Black | 154 (8.6%) | 36 (7.0%) | 1.06 | 0.74-1.50 |  |
| Asian | 313 (17.6%) | 74 (14.5%) | 1.40 | 1.08-1.81 |  |
| Mixed | 23 (1.3%) | 5 (1.0%) | 0.79 | 0.32-1.93 |  |
| Other | 41 (2.3%) | 5 (1.0%) | 0.79 | 0.32-1.93 |  |
| Ward (n = 1946) |  |  |  |  |  |
| General ward | 1500 (77.1%) | 414 (69.7%) | - | - | - |
| Respiratory unit | 235 (12.1%) | 82 (13.8%) | - | - | - |
| Critical care | 211 (10.8%) | 98 (16.5%) | - | - | - |

n = number individuals with data available; aHR = adjusted Hazard Ratios from Cox survival model adjusted for age, gender and ethnicity; CI = 95% confidence interval; p = p value of likelihood ratio test of hazard model with and without variable; ref = reference covariate.

There were 594 deaths over 99 days (11.4 deaths per person-year at risk [PY]). There were 414 deaths (12.5 deaths/PY) among admissions to general medical wards and 83 deaths (8.05 deaths/PY) among admissions to RU. Two hundred and sixteen (10%) admissions included a spell in the critical care unit (CCU), with 98 recorded deaths (8.83 deaths/PY). One hundred and seventy-seven patients (8.5%) were admitted to CCU without having been transferred from or subsequently to an RU.

The median length of stay was 5 days (IQR 2-11) for all inpatients irrespective of ward. Length of stay was longer where admissions included a spell in CCU (median 13 days [IQR 6-23]) or RU (9 days [IQR 5-15], p<0.001). A considerable tail of individuals had prolonged admissions over 30 days (4% in general wards, 5% in the RUs, and 19% in CCU; Figure 1). Two hundred and ninety-three admissions included a spell in one of the RUs, of which 248 (84.6%) are included in the subgroup analysis.

**Figure 1.**
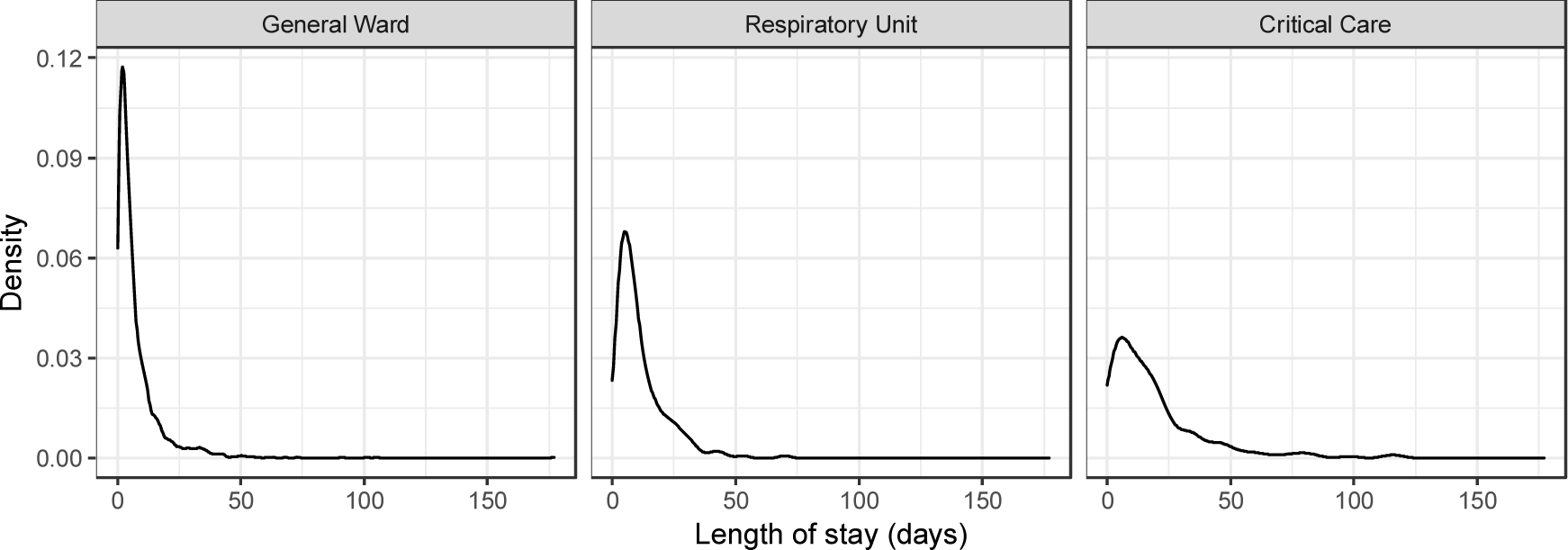
Density plot of patient length of stay by patients admitted to General Wards, the RUs, and CCU

#### 3.1.2. Survival analysis

Age, gender, and ethnicity were included as covariates in the Cox proportional hazards model (Table 1). Increasing age was associated with decreased survival probability: compared to patients younger than 60, patients aged 60-80 had 2.95 times higher rate of death, increasing to 4.44 times in those older than 80 (95% confidence interval [CI] 2.12-4.11 and 3.19-6.19 respectively). Male sex was associated with higher mortality rate than female (HR 1.30, 95% CI 1.09-1.56), as was Asian ethnicity compared to White (HR 1.40, 95% CI 1.08-1.81). No difference in survival probability was seen with Black, Mixed or other ethnicity compared to White.

### 3.2 Respiratory Units

Data were collected from 248 admissions, representing 235 individual patients. One patient was excluded due to highly missing data, and analysis is on 234 patients. Amongst 28 patients with more than one admission recorded, the average number of admissions was 2.32.

In this nested cohort, the median age was 65 years (IQR 54-80). Most patients admitted to the RUs with COVID-19 were male (66%). 76 patients died during the data collection period (9.5 deaths/PY) and 20 patients remained inpatient. Patient ethnicity was: 135 (58%) White, 46 (20%) Asian, 31 (13%) Black, 4 (2%) Mixed, and 18 (8%) Other. (Table 2)

**Table 2.** Demographic, clinical, and laboratory findings for patients admitted to the Respiratory Units

|  | Total (N=234) | Died (N=76) | Alive (N=158) | p |
| --- | --- | --- | --- | --- |
| **Age (median, IQR)** | 65 (54-80) | 79 (70-83) | 58 (50-71) | < 0.001<sup>a</sup> |
| **Male gender** | 155 (66.2%) | 48 (63.2%) | 107 (67.7%) | 0.5<sup>b</sup> |
| **Ethnicity** |  |  |  | 0.03<sup>b</sup> |
| **White** | 135 (57.7%) | 55 (72.4%) | 80 (50.6%) |  |
| **Other** | 18 (7.7%) | 3 (3.9%) | 15 (9.5%) |  |
| **Asian** | 46 (19.7%) | 12 (15.8%) | 34 (21.5%) |  |
| **Black** | 31 (13.2%) | 5 (6.6%) | 26 (16.5%) |  |
| **Mixed** | 4 (1.7%) | 1 (1.3%) | 3 (1.9%) |  |
| **CFS (n = 76)\*** |  |  |  | 0.2<sup>b</sup> |
| **<5** | 34 (44.7%) | 13 (36.1%) | 21 (52.5%) |  |
| **>=5** | 42 (55.3%) | 23 (63.9%) | 19 (47.5%) |  |
| **TEP (n = 186)** |  |  |  | < 0.001<sup>b</sup> |
| **Level 1** | 57 (30.6%) | 33 (57.9%) | 24 (18.6%) |  |
| **Level 2** | 32 (17.2%) | 17 (29.8%) | 15 (11.6%) |  |
| **Level 3** | 97 (52.2%) | 7 (12.3%) | 90 (69.8%) |  |
| **Pre-existing DM (n = 212)** | 69 (32.5%) | 24 (36.9%) | 45 (30.6%) | 0.4<sup>b</sup> |
| **Obese** | 32 (13.7%) | 8 (10.5%) | 24 (15.2%) | 0.3<sup>b</sup> |
| **CRP Adm (n = 219)** | 127 (99) | 129 (100) | 125 (99) | 0.8<sup>a</sup> |
| **Peak (n = 220)** | 213 (286) | 267 (479) | 189 (115) | 0.06<sup>a</sup> |
| **DC (n = 200)** | 97 (101) | 183 (115) | 57 (62) | < 0.001<sup>a</sup> |
| **Creat Adm (n = 228)** | 109 (82) | 117 (98) | 106 (74) | 0.3<sup>a</sup> |
| **Peak (n = 225)** | 148 (152) | 174 (165) | 137 (145) | 0.09<sup>a</sup> |
| **DC (n = 204)** | 110 (113) | 156 (166) | 88 (68) | < 0.001<sup>a</sup> |
| **ALT Adm (n = 215)** | 45 (56) | 47 (71) | 44 (48) | 0.7<sup>a</sup> |
| **Peak (n = 217)** | 98 (253) | 146 (443) | 77 (82) | 0.06<sup>a</sup> |
| **DC (n = 187)** | 66 (192) | 102 (338) | 50 (53) | 0.09<sup>b</sup> |
| **Lymph Adm (n = 233)** | 1.12 (0.97) | 1.19 (1.47) | 1.08 (0.62) | 0.04<sup>b</sup> |
| **Nadir (n = 225)** | 0.83 (0.92) | 0.90 (1.50) | 0.79 (0.46) | 0.07<sup>b</sup> |
| **DC (n = 202)** | 1.31 (1.37) | 1.18 (2.23) | 1.37 (0.69) | 0.04<sup>b</sup> |

n = number of patients with data available; Creat = Creatinine; ALT = alanine transferase; Lymph = lymphocyte count; Adm = admission result; DC = discharge result; all continuous variables expressed as mean (SD) unless otherwise specified
\* CFS only extracted for patients >65 years old
<sup>a</sup> Linear model ANOVA
<sup>b</sup> Pearson's Chi-squared tests
All tests with Bonferroni correction

Pre-existing diabetes was present in 69 patients (33%), and 32 patients had pre-existing obesity (14%). Amongst 122 patients who were admitted to the RUs above the age of 65 years, median CFS was 5 (IQR 3-6).

### 3.3 Laboratory findings

In univariable analyses, patients who survived to discharge appeared to have lower maximum recorded C-reactive protein (CRP) level during admission, lower maximum serum creatinine and lower maximum serum alanine aminotransferase (ALT) (survivors vs non-survivors, CRP: 189 mg/L vs 267 mg/L [p = 0.06], creatinine: 137 μmol/L vs 174 μmol/L [p = 0.09] and ALT: 77 IU/L vs 146 IU/L [p = 0.06]). Analysis of LDH, D-dimer, creatine kinase and ferritin were excluded due to a high degree (>50%) of missingness.

### 3.4 Non-invasive ventilation, escalation and ceilings of care

Fifty percent of patients were clinically documented as appropriate for treatment escalation to level 3 care; 17% were for NIV (level 2 care) and 33% for level 1 care. 48 patients had RU-based NIV; this consisted of CPAP in 88%. Mortality was higher in patients requiring NIV than those not (13.1 vs 8.6 deaths/PY, p=0.1), likely reflecting disease severity.

Of those who received NIV and were considered for admission to the CCU, 74% ultimately required admission (N=20/27). 46% patients receiving NIV in the RU had a trial of awake prone positioning. Awake prone positioning in patients using NIV was associated with lower mortality (8.2 vs 16.5 deaths/PY, p=0.1). Amongst patients who required NIV, mortality rate was higher amongst those considered appropriate for level 3 care compared to maximal level 2 care (3.0 vs 34.7 deaths/PY, p < 0.0001, Figure 2).

**Figure 2.**
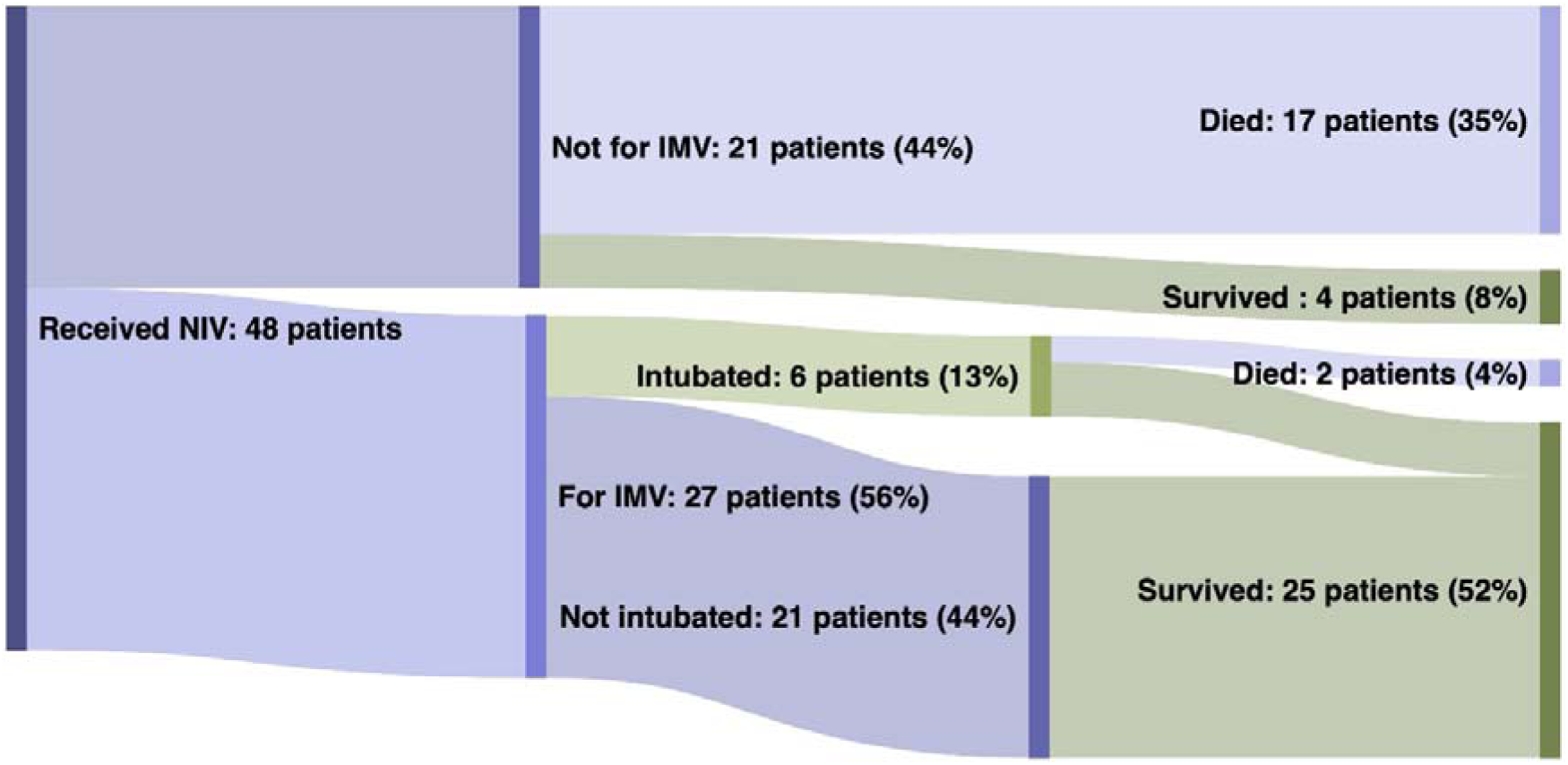
Sankey diagram showing proportion of patients who received NIV and outcomes depending on whether IMV was appropriate and if this was carried out. Percentages refer to total number of patients.

### 3.5 Radiological findings

Admission CXRs were classed as normal in 38 (17%), classical for COVID-19 in 121 (53%), demonstrating non-COVID-19 features in 46 (20%) and indeterminate in 24 (10%). There was no difference in mortality based on these CXR appearances (p = 0.4). Of the 38 patients who had normal CXR on admission, 26 had further CXRs during their admission, of which 12 were subsequently classed as classical for COVID-19. For patients with admission CXRs classical of COVID-19, mortality increased with degree of radiological severity (Cochran-Armitage, Z = −1.9641, p = 0.02).

### 3.6 Thrombosis

Eighty-four studies were performed on 66 patients to evaluate for suspected thromboembolic events, including computed tomography pulmonary angiograms (CTPA), CT head, and leg Doppler ultrasound scans. Thirteen studies were positive for thrombosis in 11 patients, representing 4.6% of patients experiencing a thromboembolic event during admission. Five patients were diagnosed with pulmonary emboli (2.1%), with 5 of 25 CTPAs performed being positive for embolism. Four patients were diagnosed with ischaemic strokes (1.7%) and three with lower limb deep vein thromboses (DVT, 1.3%). One patient had a confirmed lower limb DVT and positive CTPA.

### 3.7 Follow up data

Of the 139 patients discharged from the RUs who have been offered follow up at the end of the data collection period, 109 attended for review at 6-12 weeks after discharge. Eight patients required re-admission prior to 12-week review (three with pneumonitis, two with pulmonary embolism, two with myocardial infarction and one with stroke). All 109 patients had radiological follow up at 6 weeks; CXR appearances were persistently abnormal in 54% (N=59/109).

Ongoing symptoms were reported in 67% (N=73), with fatigue being the most common (59%); the median fatigue score was 5 (out of 10; IQR 3.5-7.0). The mean pre-admission MRC breathlessness score was 1.6 (SD 1.05); post-discharge, the mean score was 2.2 (SD 1.2). Forty-one patients (38%) had an MRC breathlessness score above their pre-admission baseline at follow up; this proportion was higher in patients without pre-existing breathlessness (45% vs 22%, p = 0.03). Other symptoms included cough (17%), memory impairment (8%) speech and language issues (7%) and hair-loss (5%).

Fifteen percent of patients had PHQ-2 scores of >3 and 6% of patients had GAD-2 scores of >3 (6%), representing a positive screen for depression and anxiety respectively. Of those who worked prior to admission, 48% felt well enough to return to work at 6 to 12-week review.

## 4. Discussion

### 4.1 Key findings

The overall mortality rate of 11.4 deaths/PY in our population is comparable to inpatient cohorts in other London hospitals.^15^ We find that increasing age, male sex and Asian ethnicity decrease survival probability in our adjusted survival analyses. Increasing CXR severity also trends with mortality.

In our RU cohort 11 patients (4.6%) had positive studies for thrombosis, in line with reports from other centres. The proportion escalated from CPAP to intubation in critical care, as well as the mortality thereafter, are comparable to unpublished data from University College London Hospital (28% and 33% respectively).^18^ Comparison of the use of ward based NIV to other busy London hospitals is difficult since many of these data are unpublished.

We report some of the first early follow up data for patients admitted with severe disease and find that a high proportion of patients remain symptomatic at 6-12 week follow up. A high proportion reported features of mood disturbance, with elevated scores on anxiety and depression screen questionnaires, and over half of those previously working had not yet returned to work and did not feel ready to do so.

### 4.2 Limitations

We present overall data, identified through clinical coding, for all patients diagnosed with suspected or confirmed COVID-19 infection. There is potential for incorrect coding of diagnoses in the case of suspected cases, resulting in both missed cases, and inclusion of individuals who did not in fact have COVID-19 in the overall cohort. For the nested RU analyses, individual patient records were reviewed by members of the respiratory team and such misclassification is less likely. Retrospective data collection using routine health records often results in incomplete data. This is evident in our study, including 19% of patients in the RU cohort who did not have a documented treatment escalation strategy; while many patients had not had biochemical parameters of interest assessed, such that these could not be included in analyses.

We report the experience of the subset of patients admitted to the RUs, as due to the limitations of data collection approaches, this was the group for which more detailed data was available. This cohort represents most unwell COVID-19 patients in our hospitals, outside of critical care. However, patients admitted directly to CCU from the Emergency Department or wards who were not stepped down to the RU were not captured. Extrapolating from the rate from overall admission data (8.5%), this represents a small but significant proportion of patients.

### 4.3 Interpretation

These analyses add to the body of research into our understanding of the disease course and the clinical outcomes of hospitalised individuals with COVID-19 in the UK. The Trust’s location and large catchment area has resulted in a larger cohort than previously published data^19^ with high deprivation indices: Barking and Dagenham has the highest index of multiple deprivation (IMD) in London.^20^ Other analyses have identified the impact of deprivation on age-standardised mortality rates from COVID-19 in the UK, with a 118% increase in death rate comparing the least deprived to the most deprived areas, a larger effect than deprivation has on all-cause mortality.^3^ We have not assessed deprivation scores on an individual level for these analyses.

The data surrounding excess coagulopathy risk associated with COVID-19 is not yet clear. Reports from Italy and China suggest a high rate of venous thromboembolism in patients with severe COVID-19 pneumonias.^16,17^ However, without a non-COVID-19 comparator group it is difficult to interpret whether this represents a higher rate of thrombosis than would be expected in hospitalised individuals with other viral pneumonias.

Increased mortality in individuals of Asian ethnicity has been reported in multiple settings.^21^ However, we do not find an association between Black ethnicity and survival in our cohort, as is reported elsewhere. In OpenSAFELY, the adjusted hazard ratio for death in individuals of Black ethnicity was 1.71 (95% CI 1.44-2.02) compared to patients of White ethnicity.^22^ This difference may reflect low power to detect a difference in mortality, given relatively fewer individuals of Black ethnicity compared to White or Asian in our population, or the effect of unmeasured and unadjusted confounding, for example by deprivation indices.

Our finding that CXR severity according to BSTI criteria trends with mortality agrees with similar data from CXR scoring systems such as the Italian Brixia score.^23^ Together with studies which correlate semi-quantitative CT scores with mortality,^24^ this adds support to the importance of radiological scores in risk stratifying outcome with COVID-19.

### 4.4 Implications

Early predictive modelling^25^ which influenced UK policy forecast that public health measures such as home isolation, quarantine and social distancing would mitigate peak healthcare demand, but strongly emphasised the possibility and large societal cost of a ‘second peak’. As public health measures are relaxed, local outbreaks have been seen in the UK and subsequent regional or national peaks remain possible. These data contribute to local resource planning and development of guidelines for such an eventuality, and again highlight the need for detailed investigation to understand any underlying causes of the excess mortality in BAME individuals.

The need for more data on recovery from COVID-19 is well recognised.^26^ Our finding of ongoing symptoms, including symptoms of mood disorders, in individuals recovering from COVID-19 has been important locally as we consider the services, including mental health and rehabilitation support, needed by our post-COVID-19 patients. So called “long COVID” is increasingly recognised, including in patients with milder disease who did not require hospitalisation, and data such as ours provide support for evidence-based guidelines for management of post-acute COVID-19.^27^

## Data Availability

Individual de-identified participant data (including data dictionaries) will be shared. Data that underlie the results reported in this article, after deidentification (text, tables, figures, and appendices). Imaging data will not be available to view. Other documents which will be available include study protocol, statistical analysis plan, analytic code. Data will be available beginning from publication and ending five years following article publication. Data will be made available to all researchers who provide a methodologically sound proposal, for their stated aims. Proposals should be directed to.

## Transparency declaration

The lead author affirms that this manuscript is an honest, accurate, and transparent account of the study being reported; that no important aspects of the study have been omitted; and that any discrepancies from the study as planned (and, if relevant, registered) have been explained.

## Contributions

Study Concept and Design All authors

Data Extraction DOC, CJC, EWS, data extraction team in acknowledgements

Data Analysis DOC, CJC

Manuscript writing All authors

Review of final submission All authors

## Acknowledgements

We would like to thank Dr Ana Otamas, Dr Apichaya Amrapala, Dr Ateeb Khan, Dr Charlotte Toms, Dr Daniel Pope, Dr Ebo Dadey, Dr Eshrina Gosal, Dr Gabi Gubo, Dr Grisma Patel, Dr Harish Shankar Kumar, Dr Helen Fielden, Dr Jonathan Ah-Chuen, Dr Kiran Desai, Dr Lani Walshaw, Dr Lennart Graebner, Dr Matthew Birch, Dr Milo Delaney, Dr Natasha O’Sullivan, Dr Pavan Kumar, Dr Rose Shendi, Dr Shivangee Sinha, Dr Stuart Innes, Dr Tess O’Neill, and Dr Tim Hill for their assistance with data collection.

## 8. Supplemental material

### 8.1 Survival analysis

Test for independence between residuals and time with scaled Schoenfeld residuals.

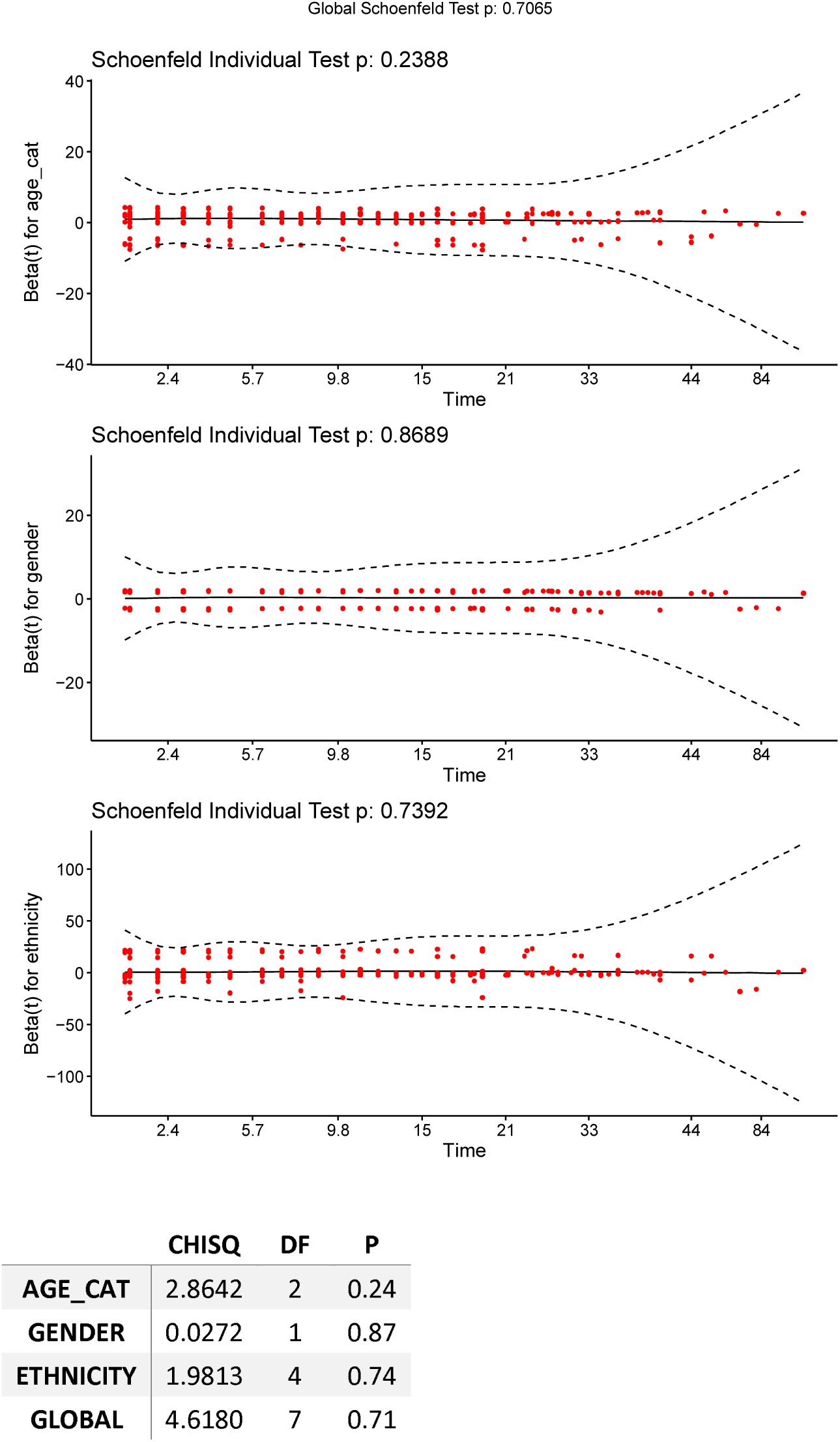

